# Successive waves in pandemic infections: physical diffusion theory and data comparisons

**DOI:** 10.1101/2020.11.03.20225235

**Authors:** Romney B Duffey

## Abstract

We establish the principal that the prediction, timing and magnitude of second and more distinct waves of infection can be based on the well - known physics and assumptions of classical diffusion theory. This model is fundamentally different from the commonly used SEIR and R_0_ fitting methods. Driven by data, we seek a working approximation for the observed orders of magnitude for the timing and rate of second and more waves. The dynamic results and characteristics are compared to the data and enable predictions of timescales and maximum expected rates where diffusive effects dominate.

The important point is this simple physical model allows understanding of the dominant processes, provides prediction estimates, and is based the solutions derived from existing, consistent and well-known physical principles. The medical system and health policy implications of such inexorable diffusive spread are that any NPI and other countermeasures deployed for and after the rapid first peak must recognize that large residual infection waves will then likely occur.

## Background : typical infection patterns and repeating “wave” phases

Since viral pandemics exhibit successive waves or peaks of infections, the key question is what physical model can explain and predict their occurrence trends and timing? The present note expands the purely numerical study by Aciola [1] by examining if the dynamic community-wide spread can be described and understood using classical diffusion theory.

To avoid confusion, the relevant fundamental physics is completely distinct from the use of the term “diffusion” by sociologists and political scientists to qualitatively explain the varying implementation of pandemic countermeasures and policies between nations [2].

The onset and subsequent progress of pandemics such as H1N1 in 1918 and Covid-19 (aka SARS-CoV-2 ) in 2020, are well known and observed to be characterized by multiple increases or “waves” of infections, peaking, declining and returning over many 100’s of days. We do not re-iterate all the well-documented features of pandemics and their behavior [see, for example 3,4,5,6], or the many and various person-to – person infection pathways and mechanisms [see e.g. 7 and 8]. We distinguish between: the dominant mechanisms of initial (direct and local transmission) rapid spreading for the first wave; and subsequently community spreading for the second and more waves (within region/country) being slowly transmitted. We postulate the first peak or “wave” is also the result of rapid initial but mainly *externally introduced* random infections as the virus attacks the first susceptible and unaware hosts. Infections grow exponentially by unconstrained random person-to-person transmission in a population without prior resistance or effective countermeasures. The initial peak (what we term here Peak 1) is reached in about 30 days, quickly spreading *between* countries/regions by which time social and non-pharmaceutical intervention (NPI) countermeasures become effective (e.g. improved hygiene, social distancing, improvised quarantines etc..) and counterbalances the growth in the infection rate. The Peak 1 wave is initial infections that grow according to initially rapid (few day) transmission and incubation timescales, subsequently declining universally according to learning theory adopting simple countermeasures [9, 10].

Zero infection numbers or rates are not achieved or achievable (as known for perennial influenza) leaving a finite probability and rate due to hidden residual infections, asymptomatic cases and/or undetected importations. This initially low level of subsequent mainly *internally introduced* or enabled community-wide spread *inside* countries is widely observed, even in island states where complete entry and viral import restrictions, mandates or controls are possible (like Hawaii, Australia and New Zealand). The virus progressively becomes fully embedded throughout the community, and largely undetected or hidden progressively continues to spread. Since the initial infective decline is competing against diffusive increase, the second wave onset only starts to be discernable or noticeable many weeks after the Peak 1 at circa 30 days. Eventually, after about 100 days, a second distinct “wave” or increase to a plateau (what we term here as Peak 2) is more clearly observed, which is also reached before some balance is achieved between increased diffusive transmission and effective countermeasures.

Even if not exactly followed by all regions, these overall data patterns, trends and surges are compelling and apparently independent of continent, culture, society or century, as shown in Figure 1 (the rates per day are normalized to the first peak and the temporal evolution measured in days exceeding 100 initial infections as a nominal start or pandemic threshold). We hypothesize that the second and subsequent waves have a fundamentally different physical spreading character, having moved from the early rapid infection phase of initial local populations dominated by rapid personal incubation timescales (3-5 days) to slower and more extensive overall community spreading (50-100 days) dominated by slower societal interactions, more cautious behaviors and residual infections ^1^. These phases may of course overlap or not be entirely distinct everywhere, so we look to data for guidance, quantification and verification.

**Figure 1.**
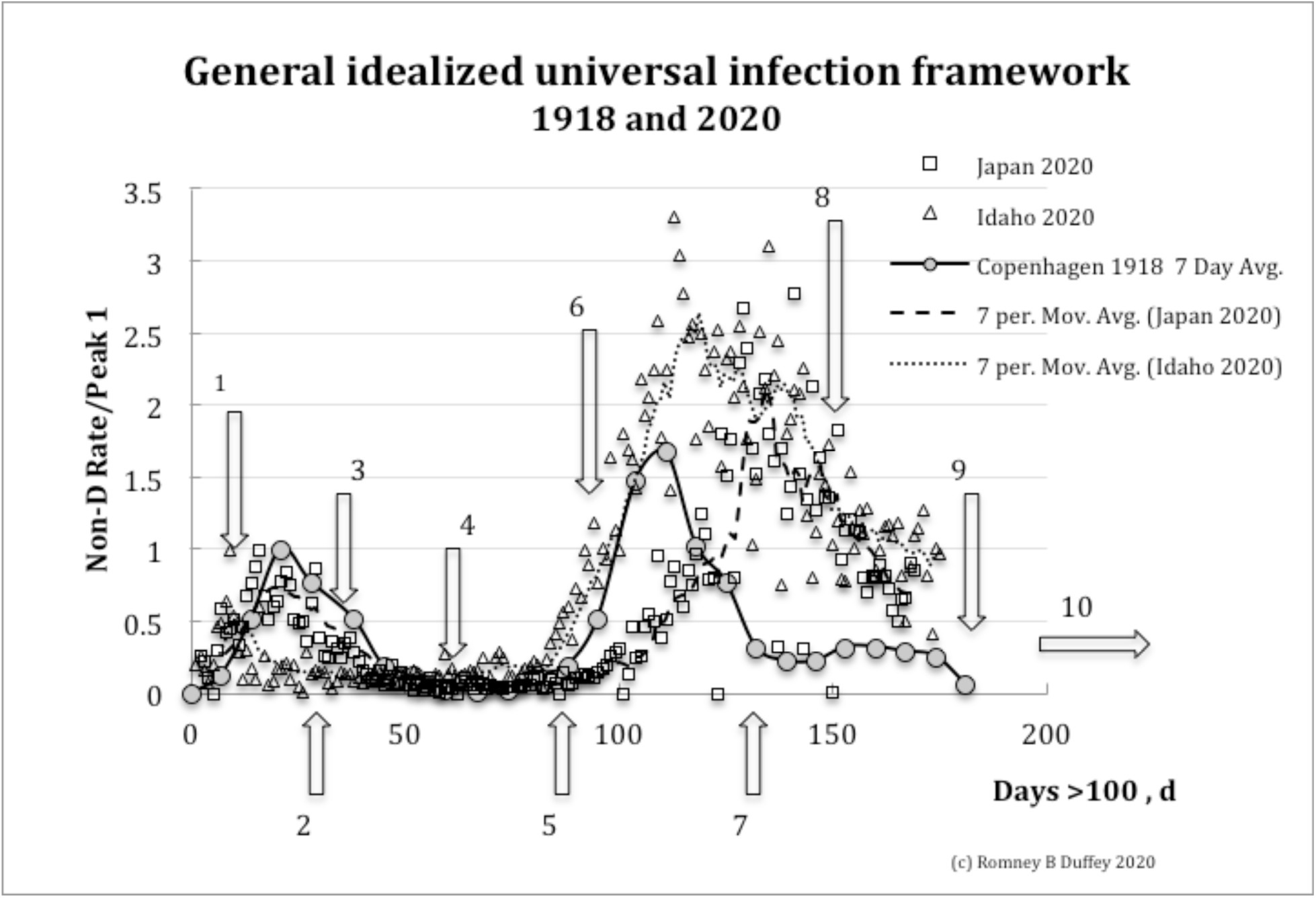
General idealized trends of pandemic infection waves over the last century, with daily infection numbers normalized to the first peak.

1. Initial random infections, n_0_, from external introduction with rapid rise (incubation limited timescale, G)
2. Exponential rise to first wave peak, n_M1_, in circa 20 - 30 days, d_M1_
3. Decline due to learning (countermeasure like awareness, hygiene, and social distancing …), k
4. Minimum achievable infection rate (or detection threshold), n_m_
5. Second “wave” apparent onset, n_02_, at circa 50-100 days (community spread limited timescale)
6. Second rise, n(d), from random inter-community spread (societally embedded)
7. Second peak or plateau, n_M2_, at about, d_M2_, circa 100+ days often larger than first peak
8. Plateau or decline due to learning, countermeasures plus human host spreading limits
9. Minimum “acceptable” or achievable rate achieved again
10. Additional waves possible (annual, seasonal, social…)

To model the physics, we postulate that after the initial peak this subsequent community-wide spreading is dominated and governed by a diffusion process, with infections seeping steadily and *inevitably* throughout the population. As first pointed out by Aciola [1] for the first peak using purely numerical solutions : “a simple diffusion model treats each individual in a population as a Brownian particle …added to this model is the incubation period of the virus and a probability of transmission of the virus if individuals are closer than a certain distance.”

Therefore, for second or other waves the problem and equation to be solved is essentially Fick’ s Law, where the net rate of change of infections at any location is proportional to the incremental infection gradient. For any infection number, n, at any time, t, conventionally:

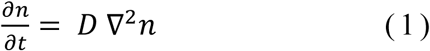

The community diffusion coefficient, D, physically represents all random person-to-person and intra-societal cross infections and is a key parameter and has to be deduced from observational data. Equation (1) is a second order differential equation, so is: (a) fundamentally more general than the first order equations used in classic R_0_ and SEIR epidemiological spread models with multiple adjustable parameters; (b) contains a very limited number of variable physically-based coefficients that can be directly tested against or fitted to data; and (c) therefore able to draw on the foundations and prior knowledge of classical physics and methods For heat and concentration diffusion. The usual SEIR and R_0_ models [11, 12 and 13] have been fitted using some seven parameters to first peak rise and decline data [e.g. 14,15] where the infection increase rate is limited by personal incubation timescales. To illuminate the physics, we seek an analytical solution to Equation (1) that is testable against publically available data.

## Method : Basic Theory, Postulates, Approximations and Derivation

Exact solutions of equation (1) are numerical while we only seek a working correlation at this time to aid our understanding. So For the second and successive “waves”, we make simplifying assumptions and reasonable approximations to enable the derivation of a basic analytical solution form For the dynamic infection number trends, the accuracy of which can be determined by later comparisons to data. Hence, even if and as countermeasures are deployed, without complete isolation or elimination the virus spreads :

a. by diffusion in any local region(s) or cities, becoming inevitably embedded in the wider community (regions of higher infection numbers naturally infect regions with lower);
b. throughout any national or regional location, all the population equally able to be randomly infected so the distribution of the virus is to first order homogenous and any and all infections are equally possible;
c. with fundamental physics limiting the extent and rate of subsequent community spreading as described by the classic Fick’s Law with diffusivity parameters averaged over the population/region /society; and,
d. for initial simplicity, being equally possible affording a simple homogenous and one - dimensional approximation, and the number of infections, n, is some fraction of the total possible, N, where usually n<<<N, depending on the overall community transmission mechanisms, societal behaviors, and countermeasure effectiveness.

For some overall societal characteristic effective transmission scale, L, after an elapsed time measured in days, d, the rate of change of infections or number counted on any day, n(d,L), Equation (1), in one dimension becomes,

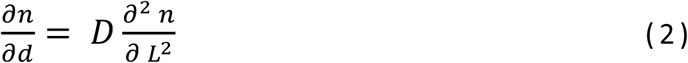

Using the usual non-dimensional formulation, 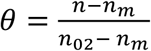, with nominal minimum achievable and second wave initial numbers, n_m_, and n_02_, respectively we have,

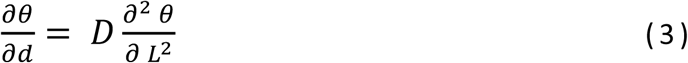

Prior physics and classic texts provides The analogous Fourier flux solution satisfying Equation (3) [1,16,17] and is the usual error function,

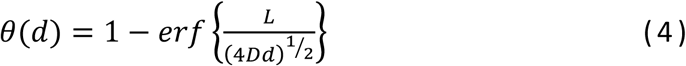

where, the term L_D_ = (4Dd)^½^ is an effective overall community viral diffusive penetration distance. To guide our thinking For present correlation and trending purposes, for the large-scale ratios, L/(4Dd)^½^, relevant to whole communities and regions during diffusion, we can retain just the first term in the series expansion [see 18 #586], so :

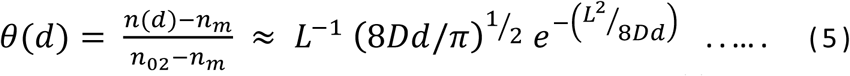

Usually, the minimum number, n_m_ << n(d) and n_02_, so 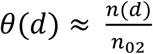. The validity, accuracy and limitations of this (or any other) approximate analytical solution must be demonstrated by data. For any chosen region this simple solution (5) has the sensible limiting conditions, *n* → *n*_*m*_ as L → ∞, *n* → *n*_02_ as d → 0, and *n* → ∞ as d → ∞ for any given diffusivity, D. After the first wave, note that we can expect to observe an initially slow square root increase in infection numbers followed by an inevitable exponential increase at longer elapsed times which is a prediction consistent with the observations (Figure 1). Writing Equation (5) in a convenient non - dimensional form, with L^*^ =L_D_/L,

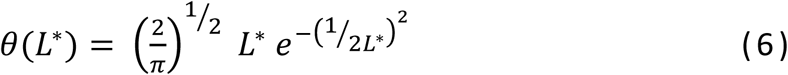

Using Equations (5) and (6) For describing the second wave requires determining just two adjustable parameters: the diffusion coefficient, D m ^2^ /d; and the assumed characteristic or effective length scale, L m, both of which of course depend on some integration/aggregation of all the exact overall societal viral transmission mechanisms (person-to-person, aerosols, crowds, contamination, random exposure …) which we simply assumed to exist.

We can then also estimate the relative size and timing, d_M2_, for attaining Peak 2 when the diffusive growth is eventually balanced, assuming the same NP I learning countermeasures are employed (e.g. social distancing, improved hygiene, public awareness etc …). For Peak 1, the decrease in the increasing rate decreases when the daily transmission and incubation increase number, n_M1_, is comparable to or balanced by countermeasures and public awareness. For some learning constant, k, representing the effect of overall countermeasures effectiveness and resilience during and after Peak 1 [10],

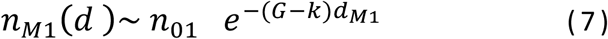

Here, the parameters for incubation growth, G, learning recovery, k, and peak day, d_M1_, are known from the Peak 1 prior data trends (Figure 1). This second wave or “curve flattening” plateau has an asymptote given by 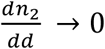, so from Equation (5), tends to flatten as, 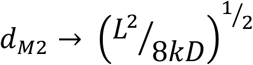, i.e. governed by the ratio of the learning reduction to diffusion increase e - folding rates. So, with countermeasures, the ratio of the Peak 1 and Peak 2 infection numbers is, for the nominal initial base or threshold numbers, n_01_ and n_02_, respectively, from equations (5) and (7),

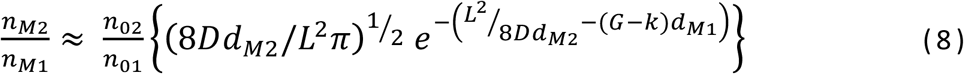

Given this approximate plateau, from equation (8) The predicted Peak 2 to Peak 1 rate size ratio is of order, 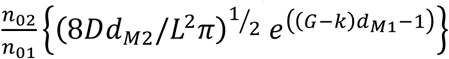, and is usually > 1 depending on effective initial learning and the ratio of the base infection numbers. Using the ratio removes any dependency on uncertainties in actual case counts, testing variations and reporting protocols.

## Results: Comparisons of theory trends to pandemic wave data

We directly examine the insights, if any, from diffusion theory using the excellent numerical data for daily infection numbers available in downloadable spreadsheet format from the open source world-in-data website^2^ (accessed ourworldindata.org/covid-cases), derived from WHO, Johns Hopkins University and other reliable sources from January 2020, including local state data from Departments of Public Health websites. As a reference origin, we adopt the initial detection/reportable threshold of n>100 total cases as a datum for the onset of significant infections, d=0, where varying this threshold has only a minor impact as initial Peak 1 usually occurs in 20 - 30 days.

The origin of the minimum achieved rate at the second wave onset, θ(d_02_), is the lowest daily infection count continuously achieved following the exponential decline from Peak 1 to normalize all subsequently increasing daily infection rates, R^*^ = θ (d)/ θ (d_0 2_). To encompass widely separated and differing societies, we performed test calculations for ongoing (as of October 15, 2020) and continuing infection data showing distinct second waves as listed in the Table 1 For UK, Italy, France, Canada, and Australia. The latter is specifically chosen as being a distinct “island state” with comparatively low (but non-zero) rates but a second wave despite extensive internal controls and border/entry restrictions.

**Table 1.** Country data with first peak and distinct second wave

| Country/region | Start d=0, 2020 | Peak 1 rate | $\theta(d_0)$day | $D \text{ m}^2/\text{d}$ | L m |
| --- | --- | --- | --- | --- | --- |
| Australia | 18 March | 715 | 81 | 0.013 | 10 |
| Canada | 12 March | 2760 | 53 | 0.012 | 7 |
| France | 5 March | 4610 | 107 | 0.014 | 10 |
| Italy | 25 February | 6560 | 100 | 0.014 | 12 |
| UK | 7 March | 5290 | 120 | 0.014 | 9 |

The comparisons to the theory are shown in Figure 2, where the fits were derived by simple comparison using the range of 0.012 <D< 0.014 m^2^ /s and 7<L<12 m values as shown in Table 1 and are largely independent of country. These fitted parameter values in Table 1 indeed are consistent with a whole society reaching a peak or plateau at 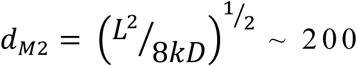 ∼ 200 days using k∼0.02/day as we previously derived for the recovery from Peak 1 in Italy and also applicable for UK and Turkey. The estimated values for the long-term diffusion coefficient in Table 1 and Figure 2 are all O(10^−2^)m^2^ /d, and for L ∼10 m, so for d∼100 days we have, L/(4Dd)^½^ ∼5, so the first term approximation for Equation (5) is reasonable).

**Figure 2.**
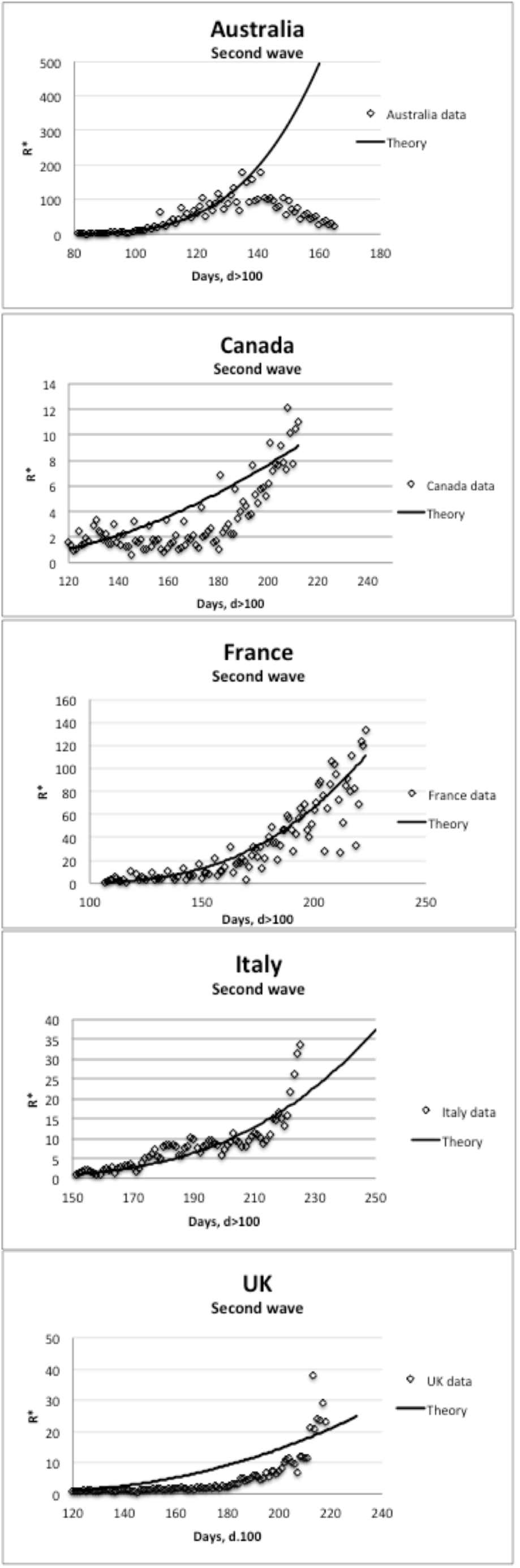
Second wave trends compared to theory.

The overall agreements with such disparate data are not perfect (Canada and UK exhibit over predictions) but the salient overall slowly growing trend and 100-200 day timescales are reasonable (especially for Australia, France and Italy) given the approximations in the theory. Capturing the steep exponential onset is also inexact at present. The Australia case with literally different boundary conditions on infection control shows clearly that Peak 2 can also be reached, while terminating the second wave growth as also observed in the classic 1918 pandemic [4,19,20].

When recovery from Peak 1 is not always complete, n_02_ >n_01_, ensuring that Peak 2 is significantly larger, which is what is actually observed. For countries already known to exhibit second wave peaks with recovery/decline, Australia actually had a peak 1 to Peak 2 infection ratio of 2.2 and Japan of 3. The still evolving USA data had a first peak of c 30,000 at 40 days which, after an initial decline, at 150 days merged into a second “wave” at 150 days almost trending to a wavy plateau or still persisting rate of some 70,0000, giving a peak ratio of circa 2.3.

Within a given society there is wide variation in *local* infection numbers and rates, so we made a more detailed “within country” analysis for the ten US states which have exhibited a distinct or discernable early Peak 1 plus evidence of already reaching Peak 2 or some plateau in daily rates after about 100+ days (accessed 15 October, 2020 at https://coronavirus.jhu.edu/data/state-timeline/new-confirmed-cases/), As shown in Table 2, there is a wide range of ratios with an average of 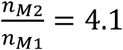, higher than the above national average. These observations can be compared to the predicted Peak 1 to Peak 2 number ratio from the diffusion model (equation (8)). Using the above observed and fitted values of d_M1_ ∼30 days, D∼0.01 m^2^ /d, L∼10 m, d_M2_ ∼100 days, (G-k) ∼0.15 /day, gives 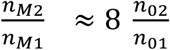, or a second Peak 2 or plateau of even an order of magnitude larger than the Peak 1 preceding it.

**Table 2.**
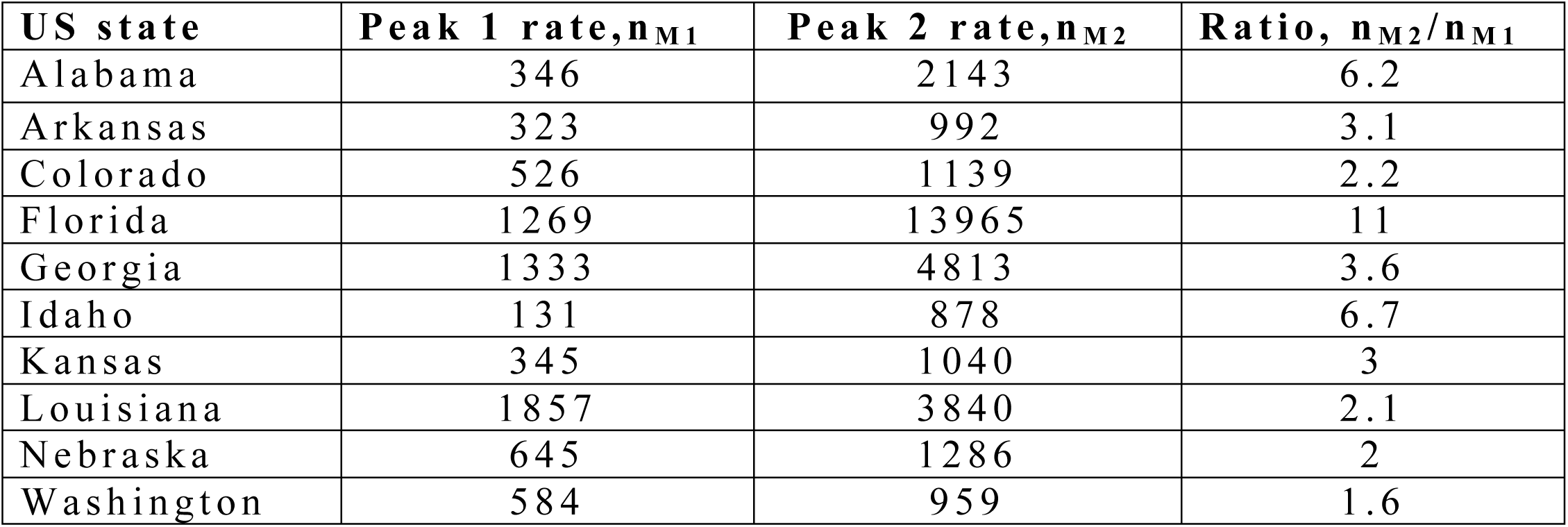
US state data with first peak and distinct second wave

| US state | Peak 1 rate, $n_{M1}$ | Peak 2 rate, $n_{M2}$ | Ratio, $n_{M2}/n_{M1}$ |
| --- | --- | --- | --- |
| Alabama | 346 | 2143 | 6.2 |
| Arkansas | 323 | 992 | 3.1 |
| Colorado | 526 | 1139 | 2.2 |
| Florida | 1269 | 13965 | 11 |
| Georgia | 1333 | 4813 | 3.6 |
| Idaho | 131 | 878 | 6.7 |
| Kansas | 345 | 1040 | 3 |
| Louisiana | 1857 | 3840 | 2.1 |
| Nebraska | 645 | 1286 | 2 |
| Washington | 584 | 959 | 1.6 |

Having made these predictions and estimates, we examined the apparently unique case of China ^3^ : (a) being the original source of the pandemic reaching Peak 1 in 26 days of 4000 cases per day by February 13 2020; (b) exhibiting a rapid decline from Peak 1 which followed learning theory and presaged global recovery trends [9]; but (c) thereafter reporting low daily infection rates (claimed to be mainly importations), with a barely discernable second wave Peak 2 reported as 276 cases per day on July 31;and (d) maintaining almost complete control of population movement and mobility, with enforced NPI mandates and personal tracking. With low and fluctuating numbers, it is difficult to uniquely pinpoint the second wave onset day, d_NO2_, and rate value, n_02_, taken here as nominally day 100 with just 2 reported cases per day, suggesting n _02_ /n _01_ ∼ 2/100 =0.02, which would give n_M2_ /n_M1_ ∼ 0.16, compared to that reported or observed of 276/4000 ∼ 0.07.

The comparison of the data to the fitted theory (Equation (5)) is shown in Figure 3 using the same D=0.01 2 m^2^ /d and L=10 m found for the Table 1 countries. This similarity suggests the fundamental physical diffusion and societal transmission processes are globally the same; and that China exhibits the same long-term embedding in the community as everywhere else.

**Figure 3.**
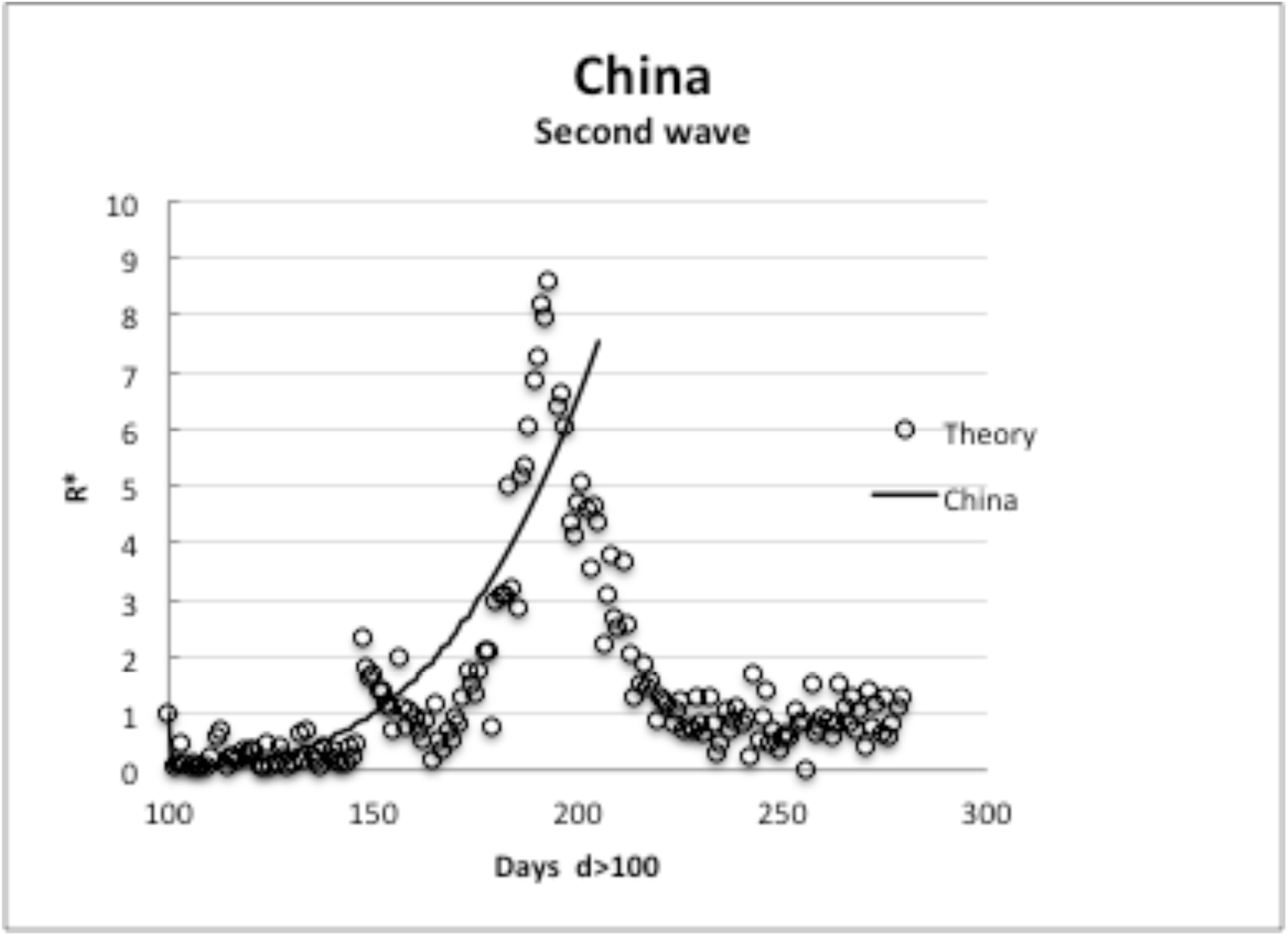
Comparison of theory to second wave data in China.

While not claiming complete accuracy as further confirmation, as of the submission date of this note, a wide range of second defined waves or plateaux after troughs following Peak 1 are on-going and have not yet really declined (e.g. current ratio examples include Austria (3), Belgium (8), Denmark (4), France (8), Germany (2.3), Indonesia (4.5), Iran (2), Israel (9.5), Italy (3), Nepal (7.6), Spain (2.3), Sweden (2), Canada (1.6), and UK (5)). These overall country ratios cover a similar range (2-12) to the US internal/regional state ratios in Table 2, and encompass the Equation (8) estimate, which surely is not a coincidence.

Presuming diffusion is the limiting phenomena, the present simple 1-D theory cannot replicate every detail for more complex scenarios but can plausibly explain as follows :

a. no discernable first peak appears because multiple numerous initially rapid infections dominate initial national counts, followed by more widespread cases over the 100’s of days of slow diffusive timescale, notably as observed in Argentina and Brazil.
b. only a slow recovery occurred after the initial rapid Peak 1, so many thousands of cases were still occurring during the diffusion masking the second wave onset which is then superimposed after about 150 days, as notably demonstrated by Russia and the USA
c. insignificant local initial infection numbers occur outside initial peak regions but increase or onset of a peak also after about 100-150 days solely due to slow progressive internal diffusive spread throughout a region, notably say for states like Alabama, Alaska,, Wisconsin and some European countries.

Beyond the scope of the present paper, these detailed trends require a more complex intra-regional analysis. An initial analysis for case (a) of no local Peak 1 but a “second wave” emerging within the community after 100 days found the same D and L parameters as for whole countries which further supports the basic diffusion concept.

Ascribing physical significance, the overall characteristic diffusive length scale of approximately 10 m is then an effective *overall* societal distance not inconsistent with realistic or feasible transmission dimensions [21]. The order of magnitude for, D, is a factor of more than 10,000 smaller than the 10^−2^-10^−3^ m^2^ /s associated or expected for local airborne atmospheric particulate or aerosol spreading mechanisms [see e.g. 21,22] reflecting the slow onset of second and other waves are simply the consequences of normal human interactions and overall social risk-taking behavior [23]. The only perfect diffusion “barrier” or so-called “circuit breaker” is complete isolation for everyone everywhere, which is not feasible and why pandemic second and more waves persist.

## Conclusions

We have made second wave size and timing predictions by adopting the dominant physics of community spreading as being diffusion limited. This simple physical concept explains the successive “waves” or recurrences of high infection numbers after 100’s of days since the first cases. Any residual, asymptomatic or undetected infections ensure the virus becomes slowly and inexorably embedded throughout the entire community after the initial rapid infection onset and partial recovery due to countermeasures.

To obtain physical insight, we have derived explicit approximate equations using the simplest homogenous one-dimensional diffusion model. Two physically-based variables, a characteristic societal length scale and an effective community wide diffusion coefficient, provide the major trends in the daily infection number trends for second waves have been fitted to the data for a wide range of countries and regions. The relative size of the second and subsequent peaks have been estimated where the peak or plateau of the second wave(s) is also limited by the relative (in)effectiveness of NPI countermeasures and societal learning effects. The policy implications of such inexorable diffusive spread must be by recognizing potentially larger infection waves will inevitably occur and planning for that eventuality. Further work is suggested to extend the physical diffusion theory concept to multiple regions and dimensions.

## Data Availability

Data publicly available via world-in-data website

https://ourworldindata.org/covid-cases

## Acknowledgments

The author is indebted all the contributors to infection data collection and reporting that have made this work possible. He also thanks Dr. E D Hughes for his important critique and suggestions; and Professors Enrico ZIo and Francesco D’Auria for their comments, encouragement and extended dialogs on this topic.

## Nomenclature

D: Effective community diffusion coefficient
d: Elapsed time in days
G: Initial characteristic infection rate
k: Learning countermeasure characteristic rate
L: Characteristic transmission societal scale
L*: Diffusive scale ratio, (4Dd /L ^2^)^½^
NPIN: on-pharmaceutical intervention
n: Number of infections in any day
R^*^: Infection number ratio, R^*^ = θ (d)/ θ (d_0_).
t: Time
θ: Non-dimensional number ratio (eq 5)
Subscripts
M1: Maximum value at Peak 1
M2: Maximum or asymptotic value of second wave
m: Minimum achieved or attainable value
0: Initial or beginning value
01: Initial value for onset of Peak 1
02: Initial value for onset of second wave

## Footnotes

1 We utilize infection rates as a leading indicator of spread, while public health of ficials usually focus on death numbers and rates which lag infections; and which fraction of infections is variable being highly dependent on the propensity, vulnerability, medical treatment and ages of the population so cannot be solely diffusivity dominated.

2 Research and data contributors listed as Hannah Ritchie, Esteban Ortiz-Ospina, Diana Beltekian, Edouard Mathieu, Joe Hasell, Bobbie Macdonald, Charlie Giattino, and Max Rose; and web development Breck Y units, Ernst van Woerden, Daniel Gavrilov, Matthieu Bergel, Shahid Ahmad, and Jason Crawford

3 This case study was suggested by Professor Francesco D’ Auria as being of particular importance in terms of the total population at risk.

## References

1. Aciola, P H, 20, Diffusion as a First Model of Spread of Viral Infection, 2003.11449v2 [physics.ed-ph] 23 May

2. Sebhatu, A, K. Wennberg, S. Arora-Jonssond, and S. Lindberg, 2020, Explaining the homogeneous diffusion of COVID-19 nonpharmaceutical interventions across heterogeneous countries, Proc NAS,117(35) 21201–21208 (accessed at pnas.org/cgi/doi/10.1073/pnas.2010625117)

3. Glezen, P W, 1996, Emerging infections: pandemic influenza, Epidemiologic Reviews, 18(1), 64–76

4. Barry, J M, “The Great Influenza”, 2005, Penguin Random House Books, New York, ISBN9780143036494

5. CDC, 2020, Coronavirus Disease 2019: learn more, US Centers for Disease Control, accessed at www.cdc.gov

6. Binti H FA, C Lau, H Nazri, DV Ligot, G Lee, C L Tan CL, et al. CoronaTracker: Worldwide COVID-19 Outbreak Data Analysis and Prediction. [Preprint]. Bull World Health Organ. E-pub: 19 March 2020. doi.org/10.2471/BLT.20.255695

7. National Academies of Sciences, Engineering, and Medicine. 2020. Airborne Transmission of SARS-CoV-2: Proceedings of a Workshop — in Brief, Washington, DC: The National Academies Press.

8. Covello, V. and Hyer, R., 2020, COVID-19: Simple Answers to Top Questions, Risk Communication Guide, Association of State and Territorial Health Officials (ASTO), July 12, 2020. Arlington, Virginia. (available at www.astho.org/COVID-19/Q-and-A/)

9. Duffey, R B and E Zio, 2020a, Analysing Recovery From Pandemics by Learning Theory: The Case of COVID-19, IEEE Engineering in Medicine and Biology Society Section, 6, pp. 110789 –110795, DOI: 10.1109/ACCESS.2020.3001344

10. Duffey, R B and E Zio, 2020b, Prediction of COVID - 19 infection, transmission and recovery rates: A new analysis and global societal comparisons, Safety Science, 129, doi.org/10.1016/j.ssci.2020.104854

11. Heesterbeek, J.A.P. A brief history of R_0_ and a recipe for its calculation, Acta Biotheor., 2002, 50, 189 –204, doi:10.1023/A:1016599411804

12. Holme, P, Masuda N (2015) The Basic Reproduction Number as a Predictor for Epidemic Outbreaks in Temporal Networks. PLoS ONE 10 (3) : e0120567. doi:10.1371/journal.pone.0120567

13. Li, J, D Blakeley, and RJ Smith, 2011, The failure of R_0_, Computational and Mathematical Methods in Medicine, Volume 2011, Article ID 527610, 17 pp, doi:10.1155/2011/527610

14. Bertuzzo, E., L Mari,D Pasetto, S Miccoli, R Casagrandi, M Gatto & A Rinaldo, 2020, The geography of COVID - 19 spread in Italy and implications for the relaxation of confinement measures, Nature Communications, 11 : 4264, 2 - 11, doi.org/10.1038/s41467-020-18050-2 (www.nature.com/naturecommunications)

15. Krishna, M V, 2020, Mathematical modeling on diffusion and control of COVID - 19, Infectious Disease Modeling, 5 (2020) 588–597, doi.org/10.1016/j.idm.2020.08.009

16. Carslaw, H C and J C Jaeger, 1959, Conduction of heat in solids, 1959, Oxford University Press

17. Mills, AF, 1992, Heat transfer, Chapter 3, ISBN 0 - 256 - 07642 - 1, Irwin, Ill., USA

18. Dwight, H B, 1968, Tables of integrals and other mathematical data, MacMillan New York.

19. Andreasen, V., C Viboud, and L Simonsen, 2008, Epidemiologic Characterization of the 1918 Influenza Pandemic Summer Wave in Copenhagen: Implications for Pandemic Control Strategies, J Infect Dis., January 15; 197(2): 270–278. doi:10.1086/524065

20. Duffey R B and E Zio, 2020c, COVID - 19 Pandemic Trend Modeling and Analysis to Support Resilience Decision - Making, Biology, 9, 156; doi:10.3390/biology9070156

21. Gorbunov, B., Aerosol particles laden with COVID - 19 travel over 30m distance, doi:10.20944/preprints202004.0546.v1

22. Korovina1, N V., I.K. Zharova, O.B. Kudryashova, S.S. Titov, 2016, Diffusion coefficient when fine aerosol media propagate in a confined volume, EPJ Web of Conferences, 110, 01029, DOI: 10.1051/epjconf/201611001029

23. Duffey, R. B., 2020, Learning about Risk: Life, death and money, ISBN –13 :9798630671134

